# DeepNeo: Deep Learning for neointimal tissue characterization using optical coherence tomography

**DOI:** 10.1101/2024.06.14.23300272

**Authors:** Valentin Koch, Olle Holmberg, Edna Blum, Ece Sancar, Alp Aytekin, Masaru Seguchi, Erion Xhepa, Jens Wiebe, Salvatore Cassese, Sebastian Kufner, Thorsten Kessler, Hendrik Sager, Felix Voll, Tobias Rheude, Tobias Lenz, Adnan Kastrati, Heribert Schunkert, Julia A. Schnabel, Michael Joner, Carsten Marr, Philipp Nicol

## Abstract

**Aims:** This study aimed to develop a deep-learning algorithm to enable a fully-automated analysis and interpretation of optical coherence tomography (OCT) pull-backs from patients after percutaneous coronary intervention (PCI).

**Methods and results:** In 1148 frames from 92 OCTs, neointima was manually classified as homogeneous, heterogenous, neoatherosclerosis, or not analyzable at quadrant level by an experienced expert. Additionally, stent and lumen contours were annotated in 90 frames to enable segmentation of lumen, stent struts and neointima. Annotated frames were used to train “DeepNeo”, a deep learning tool for prediction of neointimal tissue characteristics. Performance of DeepNeo was additionally evaluated in an animal model of neoatherosclerosis, using co-registered histopathology images as the gold-standard. DeepNeo demonstrated excellent classification performance of neointimal tissue with an overall accuracy of 75%, comparable to manual classification accuracy of two clinical experts (75%, 71%). The accurate performance of DeepNeo was confirmed in an animal model of neoatherosclerosis, where an overall accuracy of 87% was achieved. Segmentation of lumen, stent struts and neointima in human pullbacks yielded very good performance with mean Dice overlap scores of 0.99, 0.66 and 0.86.

**Conclusion:** DeepNeo is the first deep learning algorithm allowing fully automated segmentation and classification of neointimal tissue, with a performance comparable to human experts. DeepNeo might ultimately help assess vascular healing after percutaneous coronary intervention in a standardized, reliable and time-efficient manner, support therapeutic decisions and improve the detection of patients at risk of future cardiac events.

## 1 Introduction

Interventional revascularization by percutaneous coronary intervention (PCI) with stent implantation is an important treatment option for patients with obstructive coronary artery disease [1]. Despite significant advancements in the field of PCI including refinement of contemporary drug-eluting stent (DES) technology, a proportion of patients still experience stent-related events such as in-stent restenosis or stent thrombosis in the long-term [2]. The development of mature and healthy stent-covering neointima is critical to prevent these adverse events. However, delayed vascular healing can impair neointimal development and contribute to stent failure[3, 4]. Hence, immature or diseased neointima play a significant role in a substantial portion of cases with stent failure [2, 5]. Optical coherence tomography (OCT), as a high-resolution intravascular imaging modality, provides detailed visualization of the coronary vasculature and can be used to assess the mode of stent failure [1, 6, 7]. Using OCT, neointima can be visualized in vivo and characterized as either homogenous or heterogenous. Previous studies have shown that homogenous neointimal tissue has a favorable phenotype, while heterogenous neointimal tissue may be associated with de novo atherosclerosis (“neoatherosclerosis”) and a worse clinical outcome [8, 9, 10, 11, 12, 13]. Therefore, accurate detection and distinction of neointimal tissue is an important step for identifying patients at risk for stent failure. However, manual evaluation of OCT images is time-consuming and highly dependent on clinician experience, which can limit clinical availability and transferability [13]. Moreover, the visual interpretation of OCT images by clinicians in daily practice may result in missing or underestimating relevant pathological changes. Hence, more standardized approaches to OCT image analysis are necessary. Deep learning has the potential to greatly assist clinicians in accurately diagnosing patients through the analysis of medical images [14, 15]. In intravascular OCT imaging, deep learning has been successfully used to characterize native atherosclerotic lesions [16, 17]. In this study, we present the first fully-automated deep learning-based algorithm (“DeepNeo”) that enables quick and accurate automated segmentation and classification of neointimal tissue characteristics.

## 2 Methods

This study has been approved by the ethical board of Technical University of Munich, Germany in accordance with local regulations (Nr. 2023-143-S-NP). A patent application describing the technology has been filed with the European Patent Office (Application 23 179 433).

### 2.1 Data Acquisition

1148 OCT images from 92 patients who underwent clinically-indicated coronary angiography and in-stent intravascular imaging with OCT at the German Heart Center Munich were collected. OCT imaging was performed according to current guidelines [18] using a commercially available OCT system (Abbott Vascular, Santa Clara, CA). The baseline characteristics of patients are provided in Table 1.

**Table 1.**
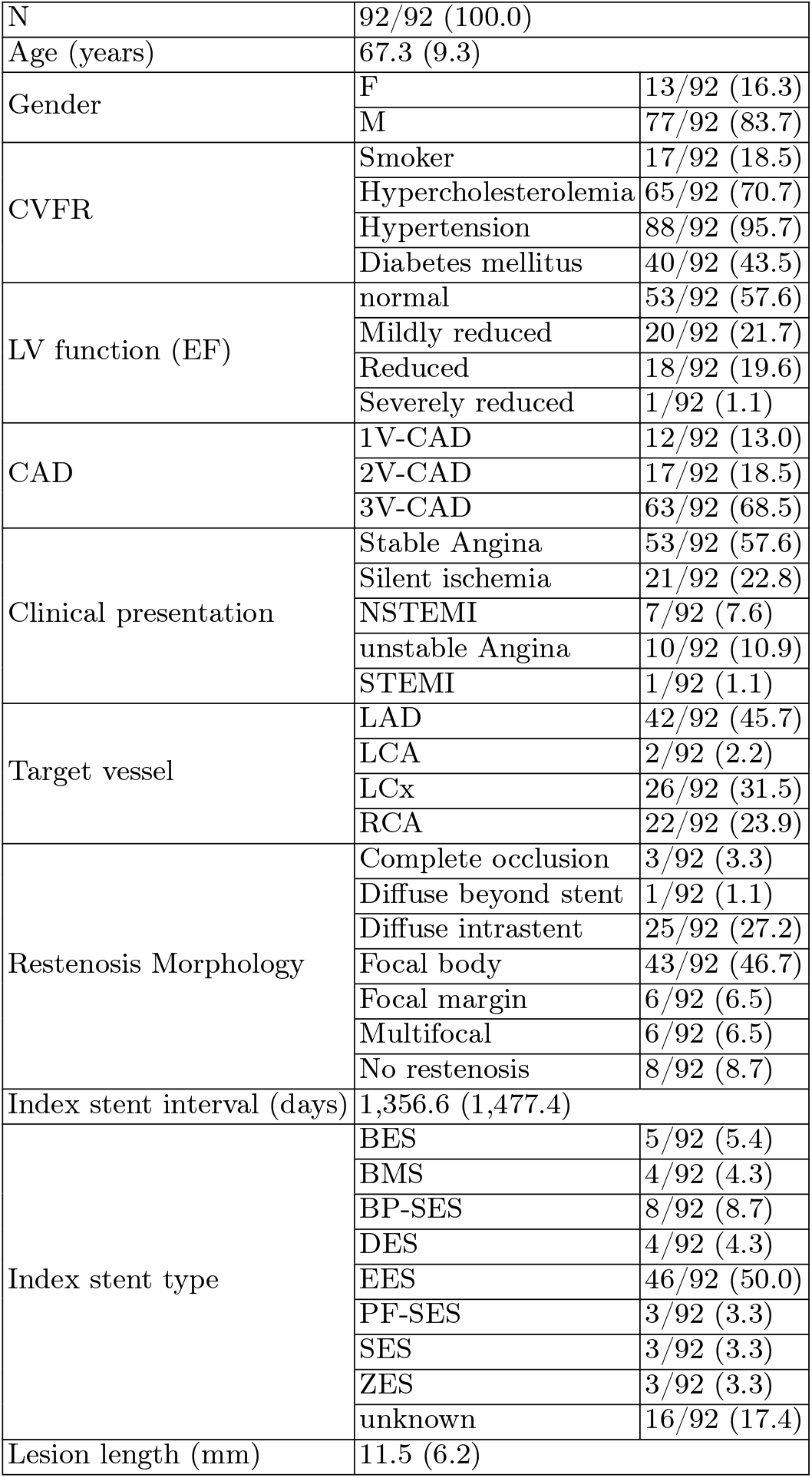
Baseline data of OCT data set Data is N/n (%) or mean (±SD)

|  |  |  |
| --- | --- | --- |
| N | 92/92 (100.0) |  |
| Age (years) | 67.3 (9.3) |  |
| Gender | F | 13/92 (16.3) |
|  | M | 77/92 (83.7) |
| CVFR | Smoker | 17/92 (18.5) |
|  | Hypercholesterolemia | 65/92 (70.7) |
|  | Hypertension | 88/92 (95.7) |
|  | Diabetes mellitus | 40/92 (43.5) |
| LV function (EF) | normal | 53/92 (57.6) |
|  | Mildly reduced | 20/92 (21.7) |
|  | Reduced | 18/92 (19.6) |
|  | Severely reduced | 1/92 (1.1) |
| CAD | 1V-CAD | 12/92 (13.0) |
|  | 2V-CAD | 17/92 (18.5) |
|  | 3V-CAD | 63/92 (68.5) |
| Clinical presentation | Stable Angina | 53/92 (57.6) |
|  | Silent ischemia | 21/92 (22.8) |
|  | NSTEMI | 7/92 (7.6) |
|  | unstable Angina | 10/92 (10.9) |
|  | STEMI | 1/92 (1.1) |
| Target vessel | LAD | 42/92 (45.7) |
|  | LCA | 2/92 (2.2) |
|  | LCx | 26/92 (31.5) |
|  | RCA | 22/92 (23.9) |
| Restenosis Morphology | Complete occlusion | 3/92 (3.3) |
|  | Diffuse beyond stent | 1/92 (1.1) |
|  | Diffuse intrastent | 25/92 (27.2) |
|  | Focal body | 43/92 (46.7) |
|  | Focal margin | 6/92 (6.5) |
|  | Multifocal | 6/92 (6.5) |
|  | No restenosis | 8/92 (8.7) |
| Index stent interval (days) | 1,356.6 (1,477.4) |  |
| Index stent type | BES | 5/92 (5.4) |
|  | BMS | 4/92 (4.3) |
|  | BP-SES | 8/92 (8.7) |
|  | DES | 4/92 (4.3) |
|  | EES | 46/92 (50.0) |
|  | PF-SES | 3/92 (3.3) |
|  | SES | 3/92 (3.3) |
|  | ZES | 3/92 (3.3) |
|  | unknown | 16/92 (17.4) |
| Lesion length (mm) | 11.5 (6.2) |  |

### 2.2 Segmentation of neointima, lumen and stent struts

Lumen contour and stent struts were manually annotated in 305 OCT frames from a subset (40 of the 92 pullbacks) using the freeware tool LabelMe ^1^ to enable automated segmentation of stent struts, lumen and neointimal area (see Figure 1 A). Segmentation allows analysis of patient characteristics such as average neointima thickness, detection of areas of uncovered stent struts, or the localization of the minimal lumen diameter in the stent. Also, segmentation masks allow the calculation of the center of the lumen, which is used to cut frames into quadrants. To assess the performance of DeepNeo for the segmentation of neointima, lumen, and stent struts, we employed a 5-fold cross-validation approach. This involved randomly dividing the dataset of 305 OCT frames into five equal parts (“folds”), with one fold used as a test set and the remaining four folds split into three training sets and one validation set. We repeated this process five times, with each fold used as the test set once. To prevent information leaks, frames from any unique patient were assigned to the same fold. The validation set was used to adjust hyperparameters that determine the model architecture and training procedure and choose the most suitable model.

**Fig. 1.**
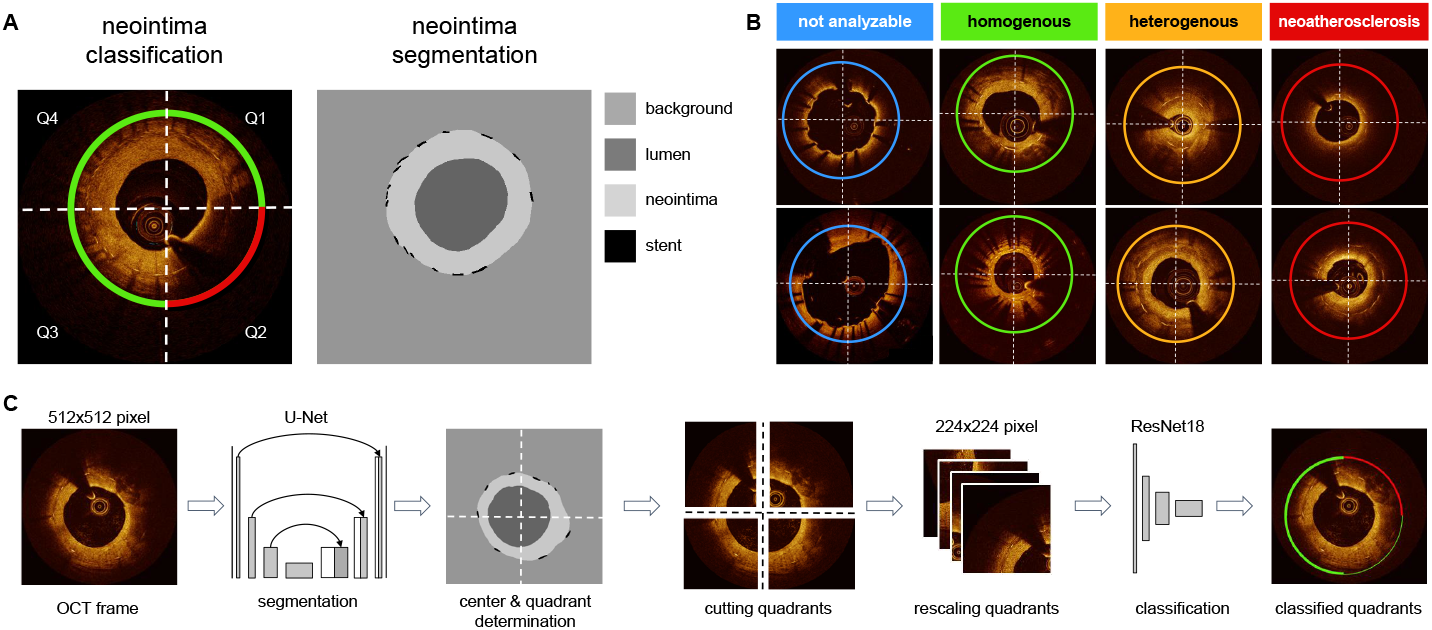
DeepNeo provides neointimal tissue segmentation and classification on quadrant level. A: OCT frames are divided into four 90° quadrants (Q1-Q4), rotating clockwise from 12 o’clock and are individually classified to one of four classes indicated by circular line color. Vessel lumen, neointima and stent struts are annotated pixelwise. B: Representative example of homogenous, heterogenous, neoatherosclerosis and not analyzable OCT frames used in the study. C: DeepNeo architecture: A frame is given as input to a U-Net to get a segmentation mask. This allows the calculation of the center of the lumen and the division of the OCT frame into 4 quadrants at the center, which are then each resized before going through the classification network. The coloured quarter-circles show the predicted class for each quadrant, line thickness indicates model certainty (thick line = high certainty).

### 2.3 Neointima classification

Manual annotation every 1 mm (every fifth frame) was performed for all pullbacks, or in adjacent suitable frames when image quality was insufficient. A total of 1148 frames from 92 pullbacks were analyzed. Neointimal tissue was classified using a quadrant-based nominal character scoring system as previously described [19]: clockwise and starting at 12 o’clock, every frame was divided into four quadrants (see Figure 1 A), with the center of the lumen as the dividing point. Each quadrant was then independently classified according to its predominant neointimal appearance into one of four classes: homogenous neointima (uniform light reflection without localized areas of stronger or weaker backscattering properties), heterogenous neointima (consisting of a focal variation of the backscattering pattern, including patterns described as “layered”), neoatherosclerosis (containing neointimal foam cells, fibroatheroma or calcifications)[20, 21] or not analyzable (quadrants with uncovered struts or side-branch openings). In quadrants with more than one tissue type, the most severe neointimal tissue type was scored. Examples of neointimal tissue types are illustrated in Figure 1 B. Expert A manually classified a total of 1148 frames (i.e., 4592 single quadrants) from 92 pullbacks. From the total of 1148 OCT frames derived from 92 pullbacks, we allocated 936 frames (originating from 66 pullbacks) to the training set. The validation set comprised 108 frames from 9 pullbacks, while the test set included 104 frames from 17 pullbacks. This test set was specifically used to assess interobserver variability and the final performance of DeepNeo, with frames being independently analyzed by experts B and C. The split was made by patient, e.g. any patient’s frames are only contained in one of the train/validation/test splits. The Fleiss Kappa score for the three independent experts was 0.654 for the test set.

### 2.4 Animal model for neointima classification

As previously published, New Zealand White rabbits underwent stent implantation in iliac arteries and repeated balloon denudation under a hypercholesterolemic diet, promoting early neoatherosclerotic lesion formation over 161 days [22]. OCT imaging and histopathological analysis of stented segments were performed using co-registration of both modalities, where OCT-frames were aligned with matching histopathology frames. The co-registration process was based on the lumen contour and the position of the stent struts in the corresponding section, as previously described [22]. Histopathology frames were divided into quadrants and scored according to the predominant tissue characteristic in each quadrant. To ensure consistency and comparability across the scoring process, we utilized a nominal character scoring system similar to that employed by DeepNeo. Specifically, a “homogeneous” score was assigned to frames demonstrating healthy neointima with a predominance of smooth muscle cells, whereas frames demonstrating infiltration with foam cells were assigned a “neoatherosclerosis” score. Frames showing deposition of fibrin, hypocellular neointima, or peristrut hemorrhage were assigned a “heterogeneous” score. It should be noted that the rabbit dataset was entirely distinct from the human dataset. DeepNeo analyzed OCT pullbacks from 12 rabbits (15 frames), and its neointimal tissue predictions were compared to the co-registered histopathology findings.

### 2.5 Algorithm architecture

We employed two deep neural networks, trained separately and combined during inference, to (i) segment lumen, stent struts, and neointima and (ii) classify the neointima in each quadrant of an OCT frame (see Figure 1C). To segment stent struts, neointima and lumen, a UNet++ was used [23]. For the classification of the quadrants, a ResNet-18 network was used [24]. To train the classification network, we divided each frame into four quadrants, using the segmentation generated by the UNet++ to determine the center of the lumen and rescaled them to a resolution of 224×224 pixels. Model calibration was achieved through temperature sharpening and fusion of the surrounding quadrants’ predictions [15]. The supplemental material provides details on augmentation techniques and training specifications for both networks.

## 3 Results

### 3.1 Segmentation of neointima, lumen and stent struts

DeepNeo achieved high accuracy in segmentation of lumen, stent struts and neointima with a Dice score of 0.99 (± 0.02), 0.66 (±0.10) and 0.86 (±0.14), respectively. The evaluation was done in a 5-fold cross-validation as described in the Materials and Methods section. The frames exhibiting inferior scoring were observed solely in regions characterized by minimal or absent neointima, thereby rendering precise annotation and prediction of the neointimal regions challenging and susceptible to marginal annotation variability (see Figure 3, low score sample).

### 3.2 Neointima classification

We compared the neointimal tissue classification performance of DeepNeo to that of clinical experts by having two additional independent specialists (expert B and expert C) manually label the test set in a blinded fashion. The labels annotated by the most experienced expert A were assumed as the ground truth and compared to the labels predicted by DeepNeo. DeepNeo achieved an accuracy of 0.75 and a macro F1-Score of 0.74, while expert B had an accuracy of 0.75 with a macro F1-Score of 0.75, and expert C had an accuracy of 0.71 with a macro F1-Score of 0.69, highlighting a high agreement of DeepNeo with the experts, which is similar to the inter-observer agreement. Figure 2 provides a comparison of manual annotations by experts A, B, and C with the automated prediction by DeepNeo. Note that frames with disagreement between experts (Fig. 2 A, B, D) resulted in lower prediction certainty (thin prediction line) compared to frames with agreement between observers (thick prediction line).

**Fig. 2.**
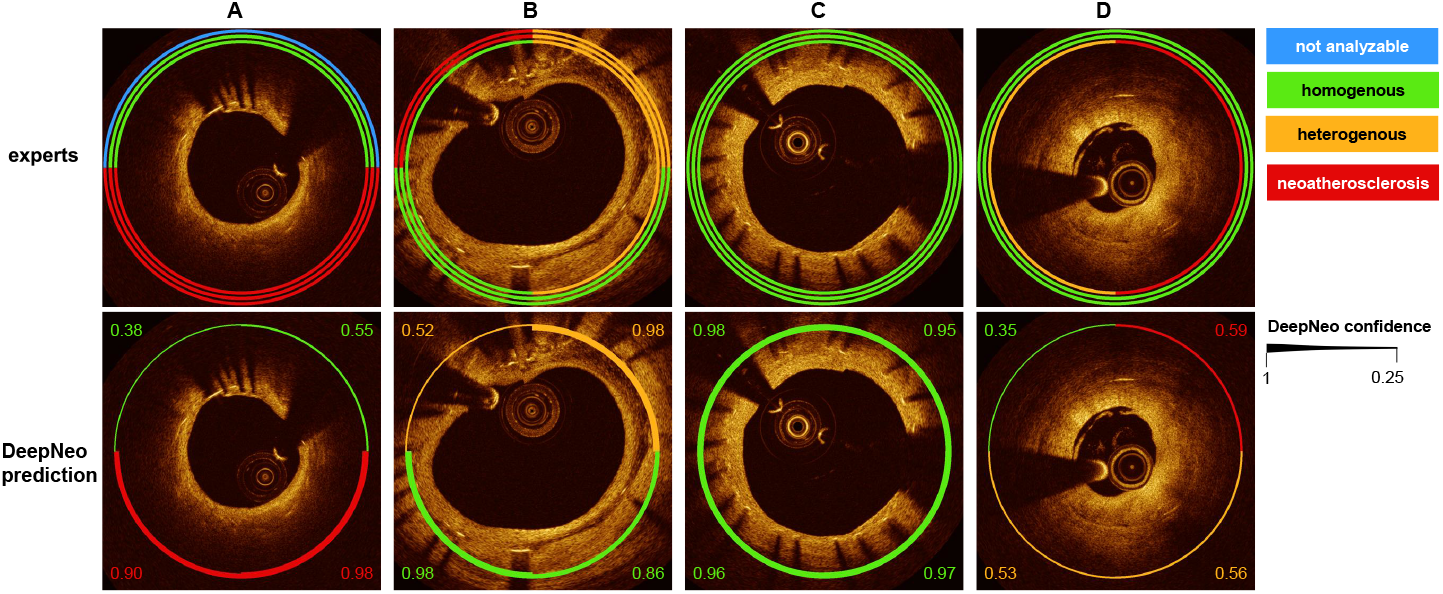
Comparison of DeepNeo predictions with manual expert classifications. Manual annotation of neointimal tissue type by three different observers is visualized by three separate circular lines. Please note that in high interobserver agreement corresponds to a high prediction certainty (c, a: quadrant 2 and 3, b: Q3) with respective thick prediction line. In contrast, interobserver disagreement corresponds to a lower certainty regarding tissue prediction, visualized by a thinner prediction line.

**Fig. 3.**
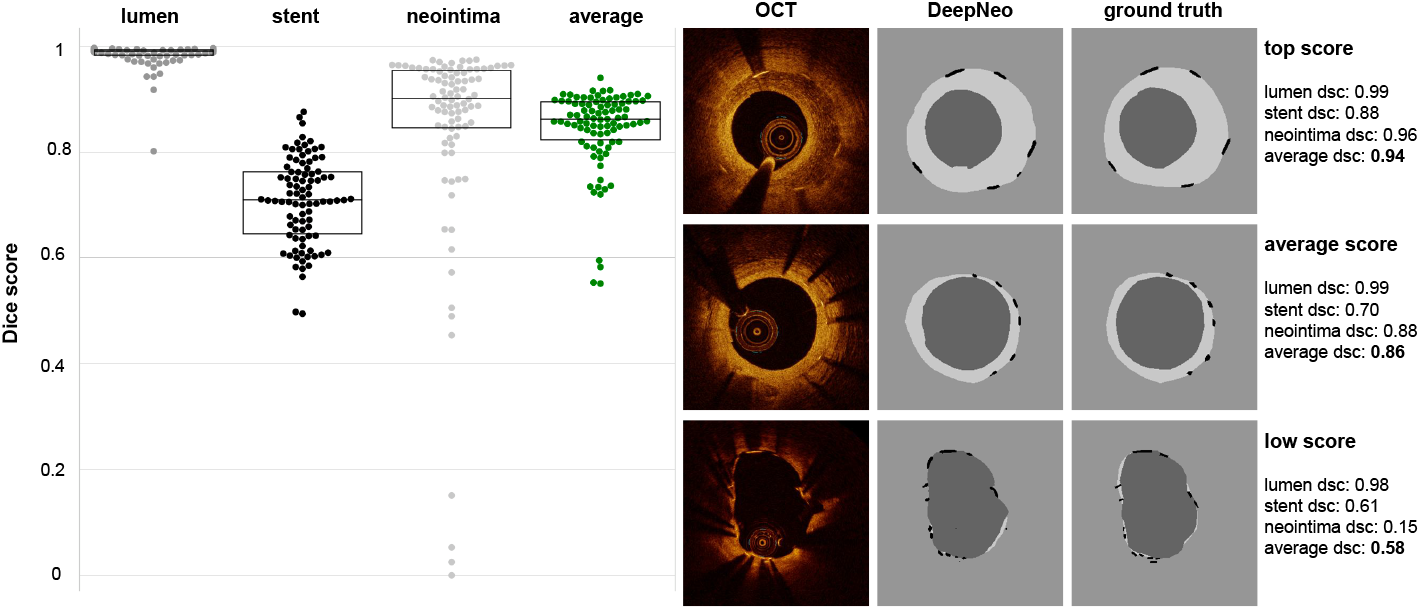
DeepNeo accurately segments lumen, stent and neointima. Beeswarm plot with boxplot, with median values (central horizontal black line), boxes extend from the 25th to the 75th percentile of scores generated by 5-fold cross-validation on 305 images from 40 patients. Good, average and low performing samples are shown with respect to the average Dice score (dsc) of an image. The Dice score is calculated as the area of overlap between labeled ground truth and prediction, ranging from 0 to 1 (0 indicating no overlap and 1 complete overlap between prediction and ground truth).

A robust correlation was observed between the model’s confidence in the predicted class and the probability of a correct prediction (Figure 4), indicating that DeepNeo is well-calibrated. This is of special importance, as it gives a notion of confidence and thus interpretability that many other algorithms lack. In supplemental figure 2 we show the need for calibration: the uncalibrated version of our model tends to be overly confident, and the correlation between the (uncalibrated) confidence and the true probability is poor.

**Fig. 4.**
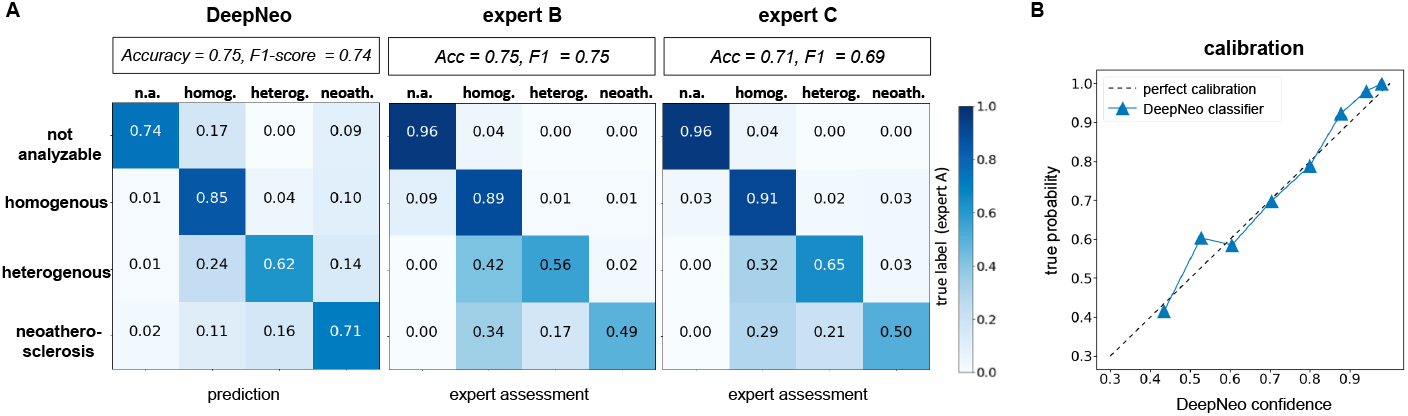
DeepNeo vs. human expert performance. Figure 3: Performance of DeepNeo and human experts. A: Confusion matrices for performance of DeepNeo and expert B and C with labels by expert A taken as ground truth. Note that automated analysis by DeepNeo is similar to the inter-expert variability. N=420 (n.a.: 23, homog.: 186, heterog.: 117, neoath.: 94). B: Calibration of DeepNeo: probability of predicted class (x-axis) vs. true probability (y-axis).

Confusion matrices shown in Figure 4 demonstrate a performance of DeepNeo similar to clinical experts. Notably, DeepNeo rarely misses diseased frames, indicating its reliability in detecting heterogenous neointima and neoatherosclerosis. Disagreement between DeepNeo and expert A, as well as inter-expert disagreement, was highest for these challenging neointimal types, with expert B and expert C sometimes leaning more towards homogenous labeling. Additional examples of DeepNeo’s automated analysis are shown in Figure 5. Analysis of failed predictions revealed that shadowing and missing stent struts were the two major sources of misclassification, as shown in Supplemental Figure 1.

**Fig. 5.**
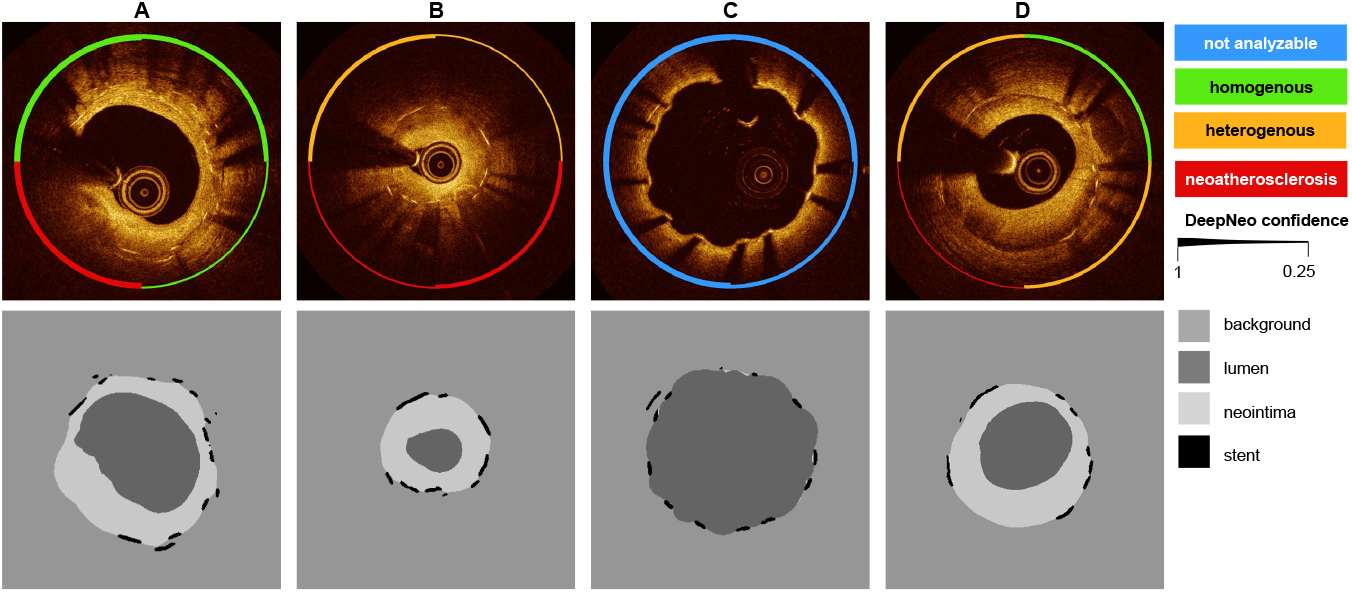
Examples of DeepNeo’s automated analysis. E Upper row: Accurate prediction of neointimal tissue characteristics on quadrant level. A: Predominant homogenous neointima with foam cells in Q3. B: Heterogenous neointima in Q1 and Q4 with foam cell infiltration in Q2 and Q3. C: No neointima present. D: Mixture of homogeneous and heterogenous neointima as well as possible neoatherosclerosis. Note the low confidence in B (Q1 and Q3) and D (Q3), reflecting the difficulty to differentiate heterogeneous neointima form neoatherosclerosis in some cases. Lower row: automated segmentation of lumen, neointima and stent struts.

### 3.3 Animal model for neointima classification

Co-registered histopathology demonstrated a high degree of concordance between DeepNeo’s predictions and the underlying tissue characteristics, as illustrated in Figure 6. DeepNeo achieved an accuracy of 0.87 and a macro F1-Score of 0.78. Specifically, DeepNeo accurately identified neointimal foam cells and fibrin deposition as neoatherosclerosis or heterogeneous, while categorizing healthy neointima with an abundance of smooth muscle cells as homogeneous. It is worth noting that DeepNeo achieved these results despite never being trained on rabbit images, demonstrating its robustness and applicability across species and acquisition setups.

**Fig. 6.**
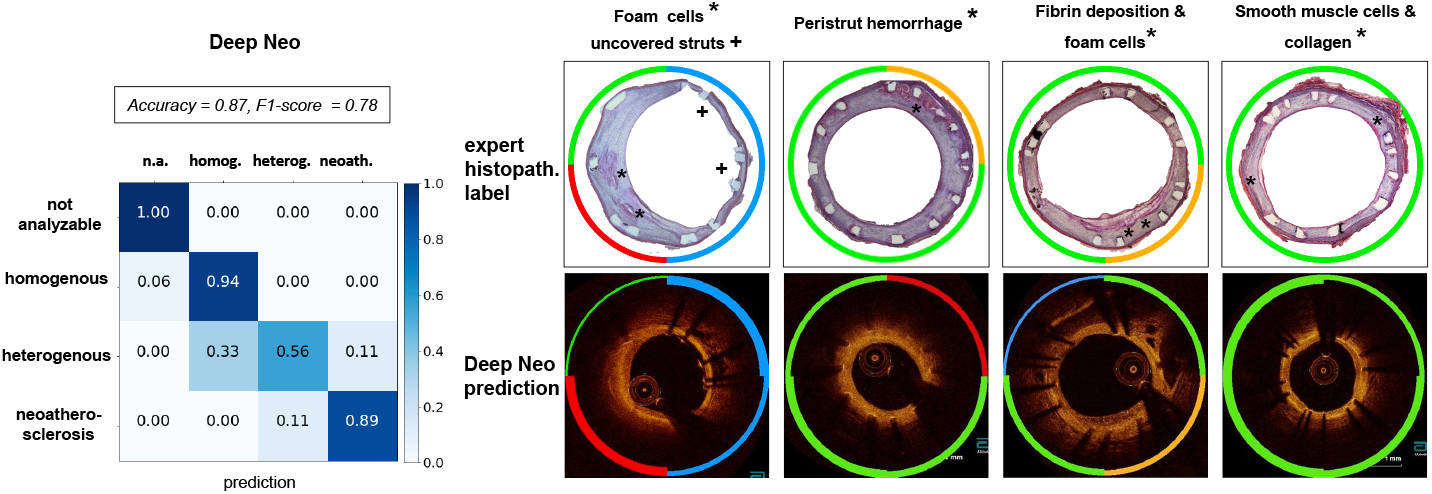
Correlation of tissue prediction by DeepNeo with histopathological findings in rabbits. A: Confusion matrix of DeepNeo with histopathological based labels. B: Representative examples from a rabbit model of neoatherosclerosis with hematoxylin-eosin (H.E.) staining, revealing underlying neointimal tissue characteristics. DeepNeo-based analysis of co-registered OCT frames showed overall good agreement between histopathological findings (marked by red asterisks) and AI-based tissue prediction.

### 3.4 Clinical cases

Figure 7 displays how DeepNeo is applied in two clinical cases of patients who underwent clinically-indicated OCT imaging after PCI at German Heart Center. Neointimal thickness and lumen radius are quantified in a standardized manner by DeepNeo, along with the neointimal tissue composition at pullback level. The visualizations provided by DeepNeo can guide clinicians to identify critical parts of the OCT pullback, which in turn enables a reliable and prompt first impression of the patients. The application of DeepNeo in these clinical cases highlights its potential in improving the standardization and efficiency of intravascular OCT imaging.

**Fig. 7.**
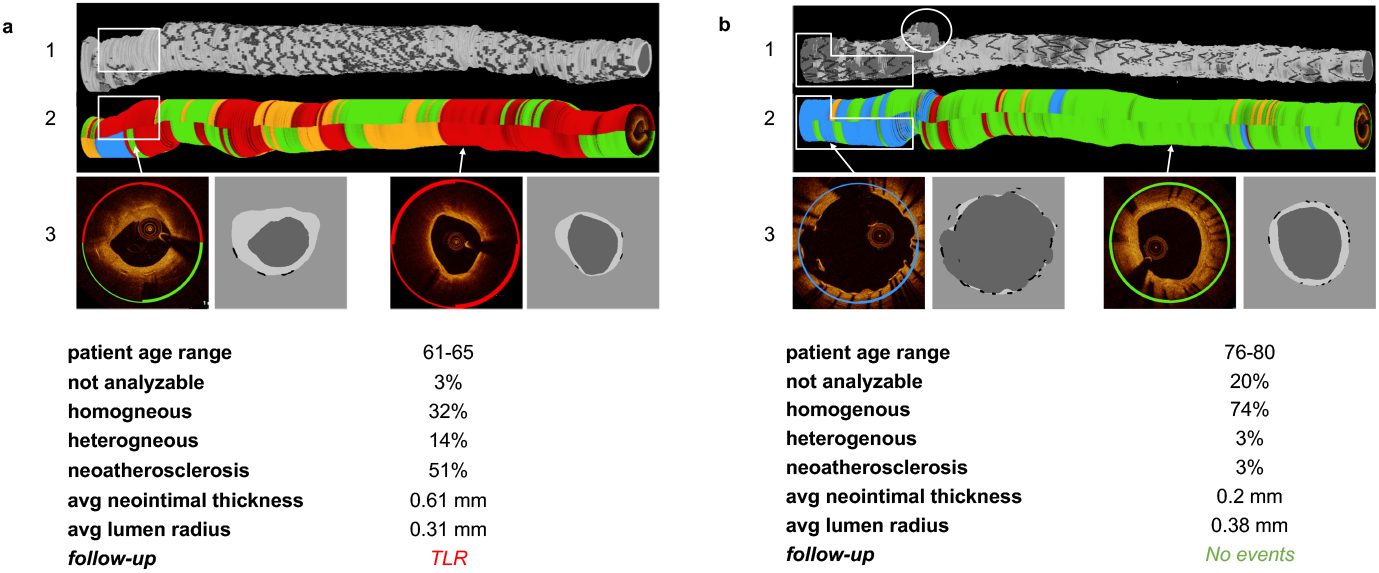
Clinical cases. 3D reconstruction of neointima, lumen and stents (1) as well as 3D reconstruction of neointimal tissue prediction (2) and sample frames (3) from two clinical cases with quantitative statistics derived from DeepNeo. A: male with PCI of RCA. OCT 12 months after PCI reveals predominately neoatherosclerotic neointima. During follow-up, the patient underwent TLR due in-stent restenosis with unstable angina. B: male with PCI of LAD. OCT 12 months after PCI reveals predominantly homogenous neointima. During follow-up, no adverse events occurred. Note how neoatherosclerosis can lead to a loss of signal leading to undetected stent struts (white box in A.1 and A.2). Note also the correct classification of uncovered stent struts as “not analyzable” (blue line in B.1 and B.2) and detection of a side-branch (white circle in B.1).

### 3.5 DeepNeo as an open-access tool

We released DeepNeo as a cloud-based open-access tool, providing a valuable resource for researchers and clinicians to rapidly analyze intravascular OCT images of stented patients. With a quick and reliable analysis time of only 2-3 minutes for a pullback, DeepNeo has the potential to greatly improve efficiency in the diagnosis. By making this freeware tool accessible to all, regardless of geographic location or financial resources, we hope to promote collaboration and accelerate progress towards better patient outcomes. The tool is built using Gradio [25], a freeware software, and hosted on Amazon Web Services Servers in Frankfurt, Germany. As demonstrated in Figure 8, DeepNeo offers a user-friendly interface that allows for easy access and analysis of intravascular OCT images. Users can simply upload their anonymized OCT pullback as a DICOM image or .zip file, with the tool providing accurate and reliable analysis. A detailed quadrant-level analysis as well as aggregated statistics over the whole pullback can be downloaded. Furthermore, the tool has the capability to determine the starting and ending points of the stent through predicted segmentation masks and subsequent postprocessing techniques. This functionality increases the degree of automation in the analysis process and reduces the need for manual intervention, ultimately decreasing the workload of clinicians and researchers.

**Fig. 8.**
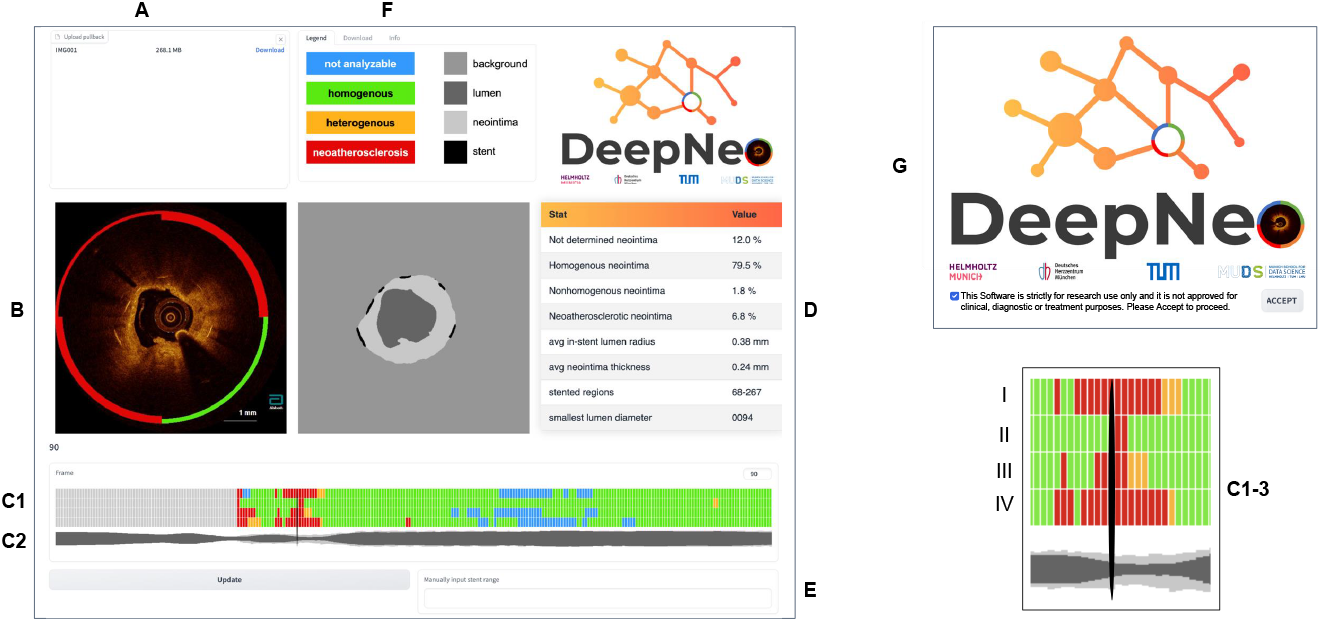
DeepNeo webinterface. The user-friendly interface is designed with several features to facilitate accurate and efficient analysis, including an upload mask (A), which allows users to upload OCT pullback images (DICOM or .zip), a visual representation of the current OCT frame with segmentation and neointima prediction, a schematic view of quadrants (C1) (top row represents quadrant I, bottom row quadrant IV) and neointima and lumen (C2) that provides a visual representation of the tissue characteristics, including a slider (C3) that enables users to move through the pullback. In addition, the interface includes a pullback analysis (D) that provides a detailed analysis of the OCT images and a manual correction feature (E) to correct beginning and end of the stent if necessary. The webtool also allows users to download a detailed analysis of their results and provides an information tab (F) for additional guidance. Users are required to accept the research-only use on the welcome page (G) before accessing the tool.

## 4 Discussion

To the best of our knowledge, DeepNeo is the first fully-automated deep learning-based algorithm for characterization of vascular healing after PCI using OCT imaging. The main components and findings are:

1. Segmentation of vessel lumen, neointimal area and stent struts, which allows further automated morphometric analysis as well as quick detection and quantification of uncovered stent struts.
2. Classification of neointimal tissue into healthy (homogenous), diseased (heterogenous or neoatherosclerosis), or not analyzable with high accuracy, comparable to performance by human observers.
3. Confirmed accuracy of prediction when using DeepNeo for analyzing OCT-pullbacks with co-registered histopathology from an animal model of neoatherosclerosis.

With millions of PCIs performed globally every year [26], there is a pressing need for effective diagnostic and therapeutic strategies to ensure optimal patient outcomes in the long term. Intravascular imaging with optical coherence tomography enables high-resolution imaging of stented lesions with detailed visualization of the neointima. Several studies have demonstrated that subjects with neointima characterized as heterogenous have a higher risk of clinical events compared to subjects with homogenous neointima [13, 27]. Additionally, heterogenous neointima might also reflect a more atherogenic milieu per se as it is associated with progression of native atherosclerosis as well [28]. Hence, heterogenous neointima following stent implantation could be regarded as a surrogate marker for poor arterial healing and adverse clinical outcome over time. Neoatherosclerosis presents an even more unstable condition [20], being detected in up to one third of drug-eluting stents [29]. Using OCT, neoatherosclerotic plaque rupture was recently identified as the major underlying cause in patients presenting with very-late stent thrombosis [30, 31]. Recently, Xhepa et al. demonstrated that detailed assessment of neointimal tissue characteristics may aid in selection of dedicated treatment strategies in patients with in-stent restenosis, showing an advantage of DES over DCB in patients with high amount of non-homogenous frames [32]. Thus, intracoronary imaging with OCT is crucial for following up on patients after PCI with stent implantation, detecting and triaging patients at higher risk of device-related events. However, interpretation of OCT images requires significant clinical expertise, and analyzing several hundred OCT frames is time-consuming and impractical in busy clinical settings. With an aging population requiring medical attention, use of deep learning-based algorithms for clinical decision support and hence reduced workload is reasonable and has already been demonstrated in different fields of medicine [33, 34]. Previous works have demonstrated the ability to segment and characterize native atherosclerotic lesions using artificial intelligence-enhanced OCT [16, 17, 35, 36]. However, to the best of our knowledge, no study so far has investigated the potential of deep learning to facilitate OCT-based characterization of neointima. We believe that DeepNeo, which allows quick and intuitive, fully-automated characterization of the underlying neointima without requiring additional human input, would be useful in following up on vulnerable patients. DeepNeo, in combination with DeepAD [17], our previously published work on the detection of native atherosclerotic lesions, provides interventional cardiologists with a useful toolbox for facilitating OCT interpretation on native as well as stented segments.

As a limitation of our study, we did not differentiate between layered neointima and heterogenous neointima, as such distinction would have reduced the sample size for each tissue class and adversely affected the performance of DeepNeo. While cross-validation would have been advantageous for classification as well, we are pleased to have 416 labels annotated by three independent experts, which we believe are sufficient for a robust evaluation. The high performance of the model on animal frames may be influenced by the limited data available for evaluation. The accuracy of DeepNeo in classifying neoatherosclerosis and heterogenous versus homogeneous neointima (71% and 62% versus 85%) may be partly explained by the increasing complexity of neointimal tissue. Homogenous neointima typically displays a simple and uniform appearance, whereas neoatherosclerosis, characterized by foam cells, calcification, or fibroatheroma, exhibits a more diverse and complex aspect that poses a challenge for accurate classification. It is worth emphasizing that a comparable reduction in performance is observed in human experts, indicating that the task of distinguishing between different types of neointimal tissue is inherently challenging. This observation suggests that the reduction in the accuracy of DeepNeo is mainly not due to a failure of the algorithm but rather a reflection of the complexity of the task. Additionally, splitting a frame into four quadrants might create ambiguous cases, such as when portions of a quadrant are more severely diseased, making classification challenging. However, it is noteworthy that misclassifications of neoatherosclerosis as heterogenous neointima or vice versa may still be considered acceptable, as both conditions are indicative of diseased tissue that requires further attention. Moreover, the identification of any diseased tissue through automated analysis can help alert clinicians to potential issues, prompting further investigation and intervention where necessary. In rare circumstances, such as inadequate contrast medium or highly atypical cases, the model may encounter difficulties; however, due to the calibrated model, those cases should result in low confidence predictions and could be flagged for further inspection. Thus, even with some degree of misclassification, DeepNeo is a valuable tool in the detection and characterization of neointimal tissue in patients after PCI.

## Data Availability

The data underlying this article will be shared on reasonable request to the corresponding authors.

## 5 Acknowledgments

**Disclosure of Interests.** T.K. is named inventor on a patent application for prevention of restenosis after angioplasty and stent implantation outside the submitted work. T.K. received lecture fees from Bayer, Abbott, Bristol-Myers-Squibb and Astra-Zeneca which are unrelated to this work. All other authors declare that they have no conflict of interest.

**Funding.** This work was supported through a scientific grant by the German Cardiac Society (Grant Number: 16/2020) and by the Helmholtz Association under the joint research school “Munich School for Data Science - MUDS”. It was also supported by the BMBF-funded de.NBI Cloud within the German Network for Bioinformatics Infrastructure (de.NBI) (031A532B, 031A533A, 031A533B, 031A534A, 031A535A, 031A537A, 031A537B, 031A537C, 031A537D, 031A538A). C.M. has received funding from the European Research Council under the European Union’s Horizon 2020 research and innovation program (grant agreement number 866411). T.K. received funding from the German Research Foundation (DFG) as part of a research project (KE 2116/4-1) and the Heisenberg program (KE 2116/5-1). Additional funding was received from the Translation & Innovation grant from Helmholtz Munich.

## SUPPLEMENT

### 1 Supplemental Material and methods

#### 1.1 Details of segmentation network

As the segmentation networks, we evaluate a standard Unet[1], Unet++[2] and DeepLabv3[3]. We observe that Unet++ performs slightly better on average than Unet, which both perform better than DeepLabv3 as can be seen in supplementary table 1. As the best model, the Unet++ with Resnet18[4] backbone is used as the segmentation network; the network implementation in Python is taken from Iakubovskii[5]. To train the Unet++, an Adam optimizer with standard parameters is used. The initial learning rate is 0.001, to regulate learning rate, a training rate scheduler by PyTorch[6] is used which reduces the learning rate by multiplying it with a factor of 0.3 if for 5 epochs the validation loss is not decreasing. The input image resolution is 512×512 with one input channel. A batch size of 8 is used with 100 training epochs. We use the Albumentations library[7] to augment images during training. Following augmentation techniques are used: Rotate, GridDistortion, ElasticTransform, HorizontalFlip, RandomBrightnessContrast, GaussNoise, RandomGamma. As the loss function, the unweighted sum of dice loss [8], cross-entropy loss and IoU loss [9] is used.

We make use of test time augmentation to increase accuracy and make the segmentation mask invariant to rotations by 90,180 and 270 degrees as well as horizontal flipping. We do so by predicting segmentation masks for not only the initial frame but also on 90, 180 and 270 degree rotated frames on the initial as well as the horizontally flipped frame. This gives a total of eight predictions per frame, and thus eight probability distributions for each pixel of a frame, which are then fused using the mean of the distributions.

**Table 1.** Comparison between segmentation performance of state-of-the art segmentation models.

|  | Lumen | Stent | Neointima | Average |
| --- | --- | --- | --- | --- |
| **Unet** | $0.986 \pm 0.025$ | $0.658 \pm 0.101$ | $0.863 \pm 0.135$ | 0.837 |
| **Unet++** | $0.986 \pm 0.021$ | $0.660 \pm 0.100$ | $0.863 \pm 0.138$ | 0.838 |
| **DeepLabv3** | $0.986 \pm 0.014$ | $0.603 \pm 0.110$ | $0.868 \pm 0.138$ | 0.820 |

#### 1.2 Details of classification network

We found that for the classification task small models suffice. Among those, a torchvision [10] Resnet18[4] showed the best performance compared to other state-of-the art networks with the same or more parameters, implemented in the torchvision library. Namely, we compare the performance to ViT-B[11] (smallest torchvision vision transformer) and Swin-T[12] (smallest torchvision Swin transformer) as can be seen in supplementary table 2. Note that for this comparison we measure the basic performance without test time augmentation, temperature sharpening, and without considering neighboring frames. As Resnet18 showed the best performance, it was selected as the network for DeepNeo.

**Table 2.**
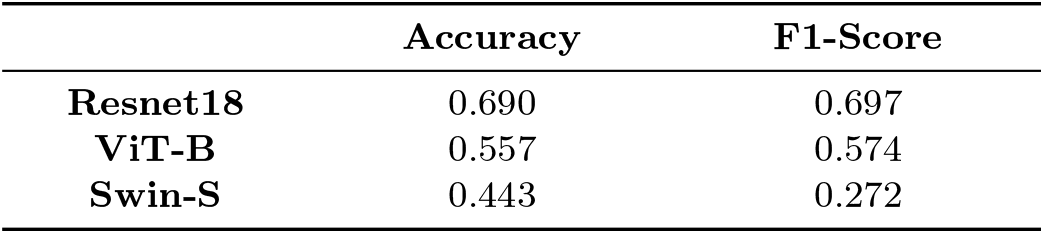
Comparison between classification performance of state-of-the art classification models.

To train the Resnet18, an AdamW optimizer with standard parameters is used [2]. The initial learning rate is 0.001, to regulate learning rate, a training rate scheduler by PyTorch [6] is used which reduces the learning rate by multiplying it with a factor of 0.3 if for 5 epochs the validation loss is not decreasing. The input image resolution is 224×224 with one input channel. A batch size of 32 is used with 100 training epochs. We use the Albumentations library[7] to augment images during training. Following augmentation techniques are used: GridDistortion, ElasticTransform, HorizontalFlip, RandomBrightnessContrast, GaussNoise, RandomGamma. As the loss function, cross-entropy loss is used. We also make use of test time augmentation here in a similar fashion as in the segmentation network: The same test-time augmentations are also used to generate eight class predictions per quadrant. The resulting distributions are fused using temperature sharpening and then normalizing, which yields slightly better results compared to mean aggregation and produces more meaningful confidence scores [13]. To account for spatial dependencies in a straightforward manner, in a final step we average the predictions over the surrounding quadrants:

For each quadrant *k* in a frame *n*, the final distribution *d*^final^ is computed:

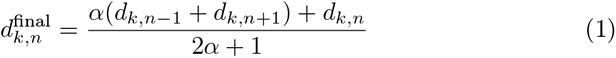

If frame *n* + 1 or *n −* 1 does not exist or is outside the stented region of the pullback, the corresponding distribution gets removed in the above equation. For example, the equation would change to:

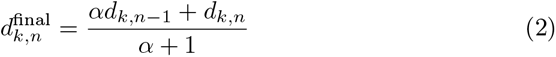

without frame *n* + 1.

The best value for was determined via grid search on the validation set and ist set to 0.75. Accounting for surrounding frames in this manner yielded a 5.7% improvement in accuracy on the test set. The resulting confidences are well calibrated as shown in Figure 4.

**Fig. S1.**
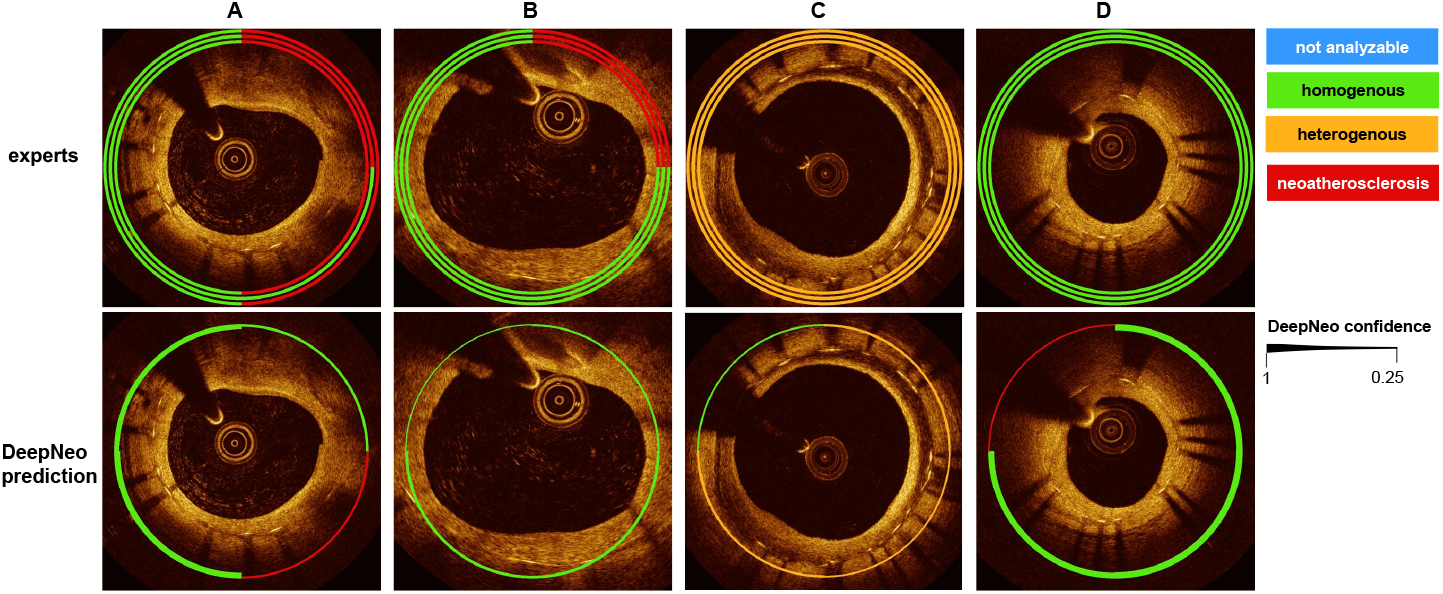
Examples for prediction failure by DeepNeo. Upper row: manual annotation by three independent observers marked by three separate lines. Lower row: automated prediction by DeepNeo. A and B: Q1 is misclassified as homogenous. C: Q4 is misclassified as homogenous. D: Q4 is misclassified as neoatherosclerosis.

**Fig. S2.**
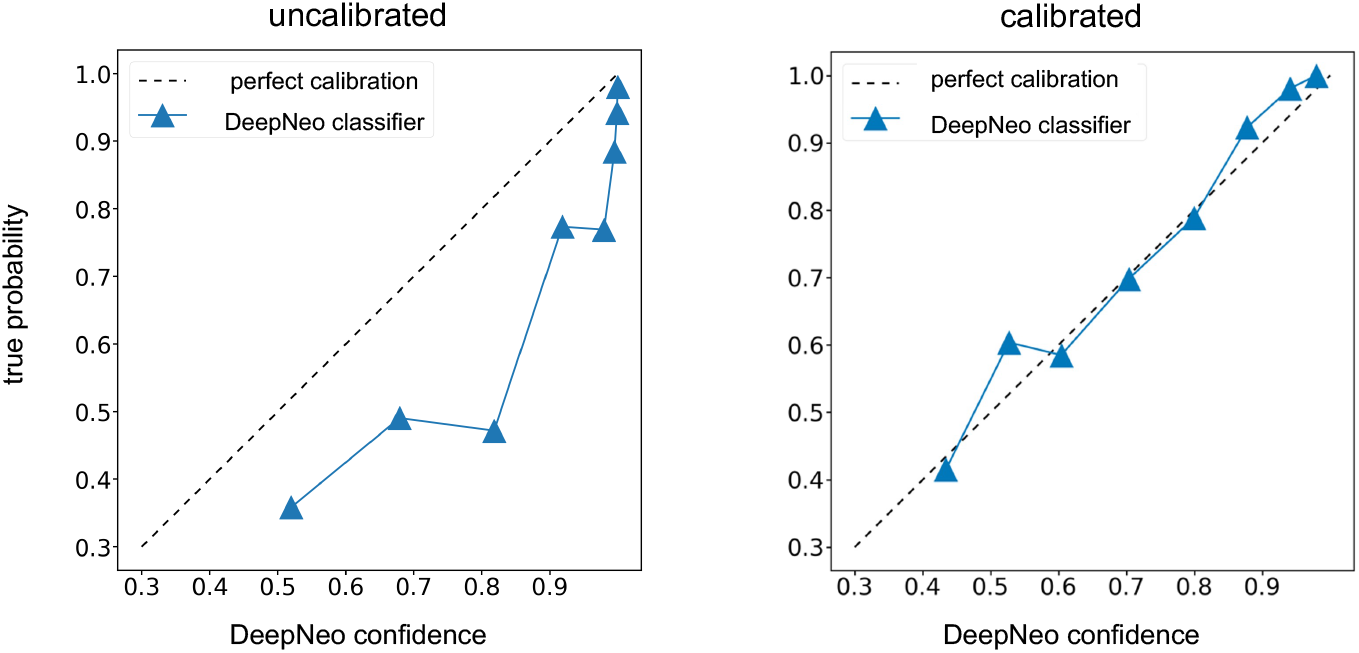
Model calibration improves correlation between confidence and true probability. Uncalibrated model (left) vs calibrated model used in DeepNeo (right).

## Footnotes

1 labelme.csail.mit.edu/Release3.0

